# Conduction system pacing is superior to reduce the new-onset atrial fibrillation risk compared with right ventricular pacing: insights from pooled clinical evidence

**DOI:** 10.1101/2024.02.20.24303113

**Authors:** Feng Li, You Zhang, Si-Liang Peng, Meng-Chao Jin, Chi Geng, Venkatesh Ravi, Parikshit S. Sharma, Pugazhendhi Vijayaraman, Hui Li

## Abstract

**Background:** Conduction system pacing (CSP) has been reported to improve clinical outcomes in comparison of right ventricular pacing (RVP). However, the performance between CSP and RVP on the risk of new-onset atrial fibrillation (AF) remains elusive.

**Methods:** Four online databases were systematically searched up to December 1^st^ 2023. Studies comprising the rate/risk of new-onset AF between CSP and RVP group were included. Subgroup analysis was performed to screen the potential determinants for the new-onset AF risk for CSP therapy. Moreover, the pooled risk of new-onset AF based on ventricular pacing burden (Vp) between CSP and RVP group were evaluated.

**Results:** A total of five studies including 1,491 patients requiring pacing therapy were eligible. The pooled new-onset AF rates for CSP and RVP group were 0.09 and 0.26, respectively. Compared with RVP group, CSP group showed a lower pooled risk (risk ratio [RR] 0.38, *P*=0.000) and adjusted risk (hazard ratio [HR] 0.33, *P*=0.000) of new-onset AF. Meanwhile, a significant intervention-covariate interaction for the adjusted risk of new-onset AF between CSP and RVP group was identified with Vp < 20% and Vp ≥ 20%.

**Conclusions:** Our study suggests that CSP is superior to reduce the new-onset atrial fibrillation risk compared with RVP. The Vp ≥ 20% may be the key determinant on the lower risk of new-onset AF with CSP therapy.

## 1. Introduction

Right ventricular pacing (RVP), including right ventricular apex pacing and right ventricular septal pacing, has been widely utilized for patients with symptomatic bradycardia owing to multiple advantages, including relatively easy implantation, satisfying efficacy, and operation safety[1, 2]. Whereas, concerns have arisen with the increasing evidence on RVP. Accumulated studies suggest that RVP is significantly associated with the ventricular electromechanical dyssynchrony, ultimately causing to increased atrial arrhythmias, heart failure, and mortality[3, 4]. Therefore, establishment of an alternative pacing strategy for decreasing the risk of new-onset AF after pacing therapy is necessary.

Conduction system pacing (CSP), including His bundle pacing and left bundle branch pacing (LBBP), or left bundle branch area pacing (LBBaP), is a promising physiological pacing strategy with the intriguing clinical outcomes, incluing reduction, even reversal of the adverse clinical outcomes related to RVP[5, 6]. Our recent published meta-analysis on the effects of CSP in patients of heart failure indicated that CSP showed a more effective performance on the heart failure patients than conventional pacing therapy, which further indicated that CSP is a superior pacing strategy to achieve a better clinical prognosis[7].

New-onset atrial fibrillation (AF) is a concern with pacing therapy due to its risk of progressing into persistent AF, increasing stroke/systolic embolism, left ventricular dysfunction, as well as impaired quality of life. Accumulated evidence demonstrated that RVP was significantly with the occurrence of new-onset AF. Whereas, evidence on the advantage of CSP versus RVP on the risk of new-onset AF remains elusive on account of limitation of small cohort of patients, lack of long-term follow-up, and a relatively high technical threshold with CSP. Therefore, we performed this registered study to explore if CSP is superior to reduce the new-onset atrial fibrillation risk compared with RVP.

## 2. Methods

### 2.1 Study design

This study was performed following the preferred reporting items for reviews and the PRISMA guidelines (**Supplementary PRISMA Checklist**). The official protocol was registered on the PROSPERO database (CRD42023492551).

### 2.2 Search strategy

Two reviewers (Feng Li and You Zhang) independently searched for four databases, including PubMed, Cochrane Library, Web of Science and Embase from the establishment of the online databases up to 1 December 2023. The searching keywords mainly included “conduction system pacing”, “CSP”, “His pacing”, “Hisian area pacing”, “His bundle pacing”, “left bundle branch pacing”, “LBBP”, “left bundle branch area pacing”, “LBBaP”, “right ventricular pacing”, “RVP”, “right ventricular apex pacing”, “right ventricular septal pacing”, “atrial fibrillation”, “new-onset atrial fibrillation”, “new-onset AF” and “new-onset atrial high-rate episodes”. The search strategy was presented in the **Supplementary Text 1**. Trials comprising the rate or risk of new-onset AF between CSP and RVP group were included. A manual search was also conducted to identify the potential publications. Moreover, the relevant corresponding authors were contacted to acquire the missing data related outcomes in their publications.

### 2.3 Study selection

Two reviewers (Feng Li and Si-Liang Peng) independently reviewed full texts for screening the eligible studies, respectively. The eligible study would be determined if inclusion criteria were met: (1) RCT and observational studies. (2) studies reporting the incidence rate of new-onset AF or (adjusted) risk ratio/hazard ratio of new-onset AF between CSP and RVP were included. (3) studies with the most data of multiple publications for the same study. Review articles, editorials, case reports, animal studies, and studies without original data were excluded. A third reviewer (Hui Li) discussed and resolved the potential disagreements on the eligibility.

### 2.4 Data extraction and quality assessment

Two researchers (You Zhang and Meng-Chao Jin), respectively, acquired the data from all eligible studies. Any controversies were consulted and resolved by a third one (Hui Li). First, we documented the eligible study characteristics, including publication year, the first author, study design, subjects type, pacing type of CSP, the sample size in the CSP and RVP group, and follow-up. Then, the study demographic and clinical characteristics, as well as adjusted condounders were also recorded.

A appraisal tool for observational studies was applied by two independent researchers (Si-Liang Peng and Meng-Chao Jin) to evaluate the quality of the eligible studies. In this study, the Newcastle-Ottawa Quality Assessment Scale (NOS), including three domains with nine points, was used. The study quality was divided into low quality (the total score < 6) and moderate-to-high quality (the total score ≥ 6). Any potential controversies were resolved by a third one (Hui Li).

### 2.5 Statistical analysis

In this study, the pooled rates of the events (ratio of the number of events to patient number) and 95% confidence interval (CI) were calculated for the new-onset AF with CSP and RVP, respectively. The pooled risk ratio (RR) and corresponding 95%CI were calculated for the risk of new-onset AF between CSP and RVP. The pooled hazard risk (HR) and corresponding 95%CI were calculated for the adjusted risk of new-onset AF between CSP and RVP, as well as the adjusted risk of new-onset AF based on the Vp. The Stata 16.0 was used for analyses. Statistically significant was defined as *P* < 0.05.

We utilized the I-squared (I^2^) and chi-squared test to quantify and evaluate the statistical heterogeneity of studies. If the I^2^ value was < 50% and/or *P* ≥ 0.05 for the chi-squared test, we defined that the between-study heterogeneity was substantial, and a fixed-effect model would be performed. Otherwise, a random-effect model was performed. Sensitivity analysis was conducted by sequentially omitting one study at a time to to evaluate the effect of a single study on the overall risk. Potential publication bias was also assessed using Egger’s test.

Additionally, subgroup analysis was conducted to screen the sources of heterogeneity, as well as the potential determinants for the new-onset AF between CSP and RVP group. Based on the study characteristics, previously reported factors and other potential factors, a total of five subgroup factors were screened, including indication of implant (SND and AVB vs. AVB), pacing type of CSP (LBBaP vs. His pacing), sample size (> 100 vs. ≤ 100), follow-up time (> 24months vs. ≤ 24months), and new-onset AF type (Clinical AF vs. Subclinical AF). Important, we also evaluated the adjusted new-onset AF risk between CSP and RVP group according to Vp (including Vp ≥ 20% vs. Vp < 20%, and Vp ≥ 40% vs. Vp < 40%).

## 3. Results

### 3.1 Study selection and quality assessment

A total of five observational studies[8–12] with 1,491 patients requiring pacing therapy (672 and 819 patients in CSP and RVP group, respectively) were eligible. The selection flowchart according to the PRISMA 2020 flow diagram (http://prisma-statement.org/prismastatement/flowdiagram.aspx) was presented in **Figure 1**. The study demographic and clinical characteristics, as well as adjusted condounders of the eligible studies were showed in **Table 1**.

**Figure 1.**
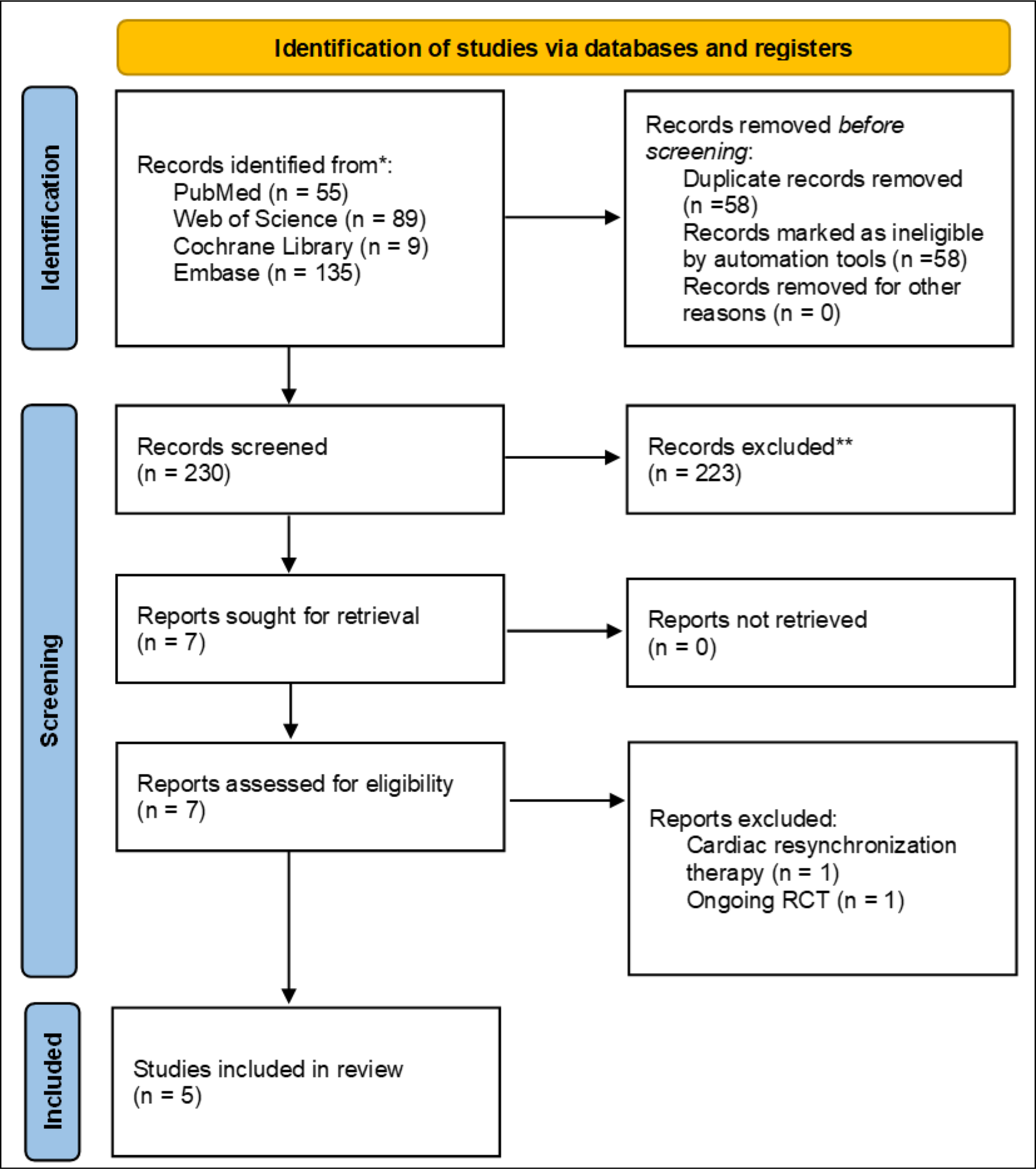
The flowchart for selecting the eligible studies.

**Table 1.**
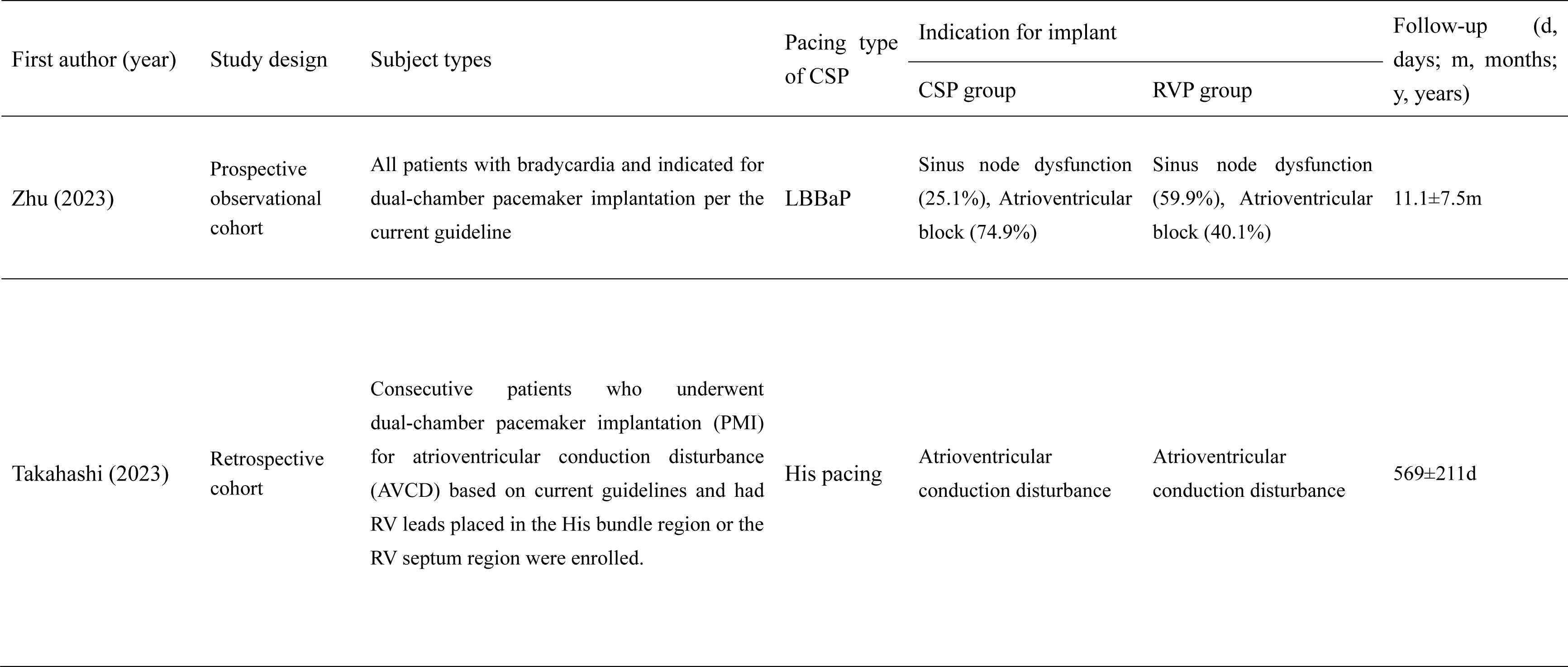
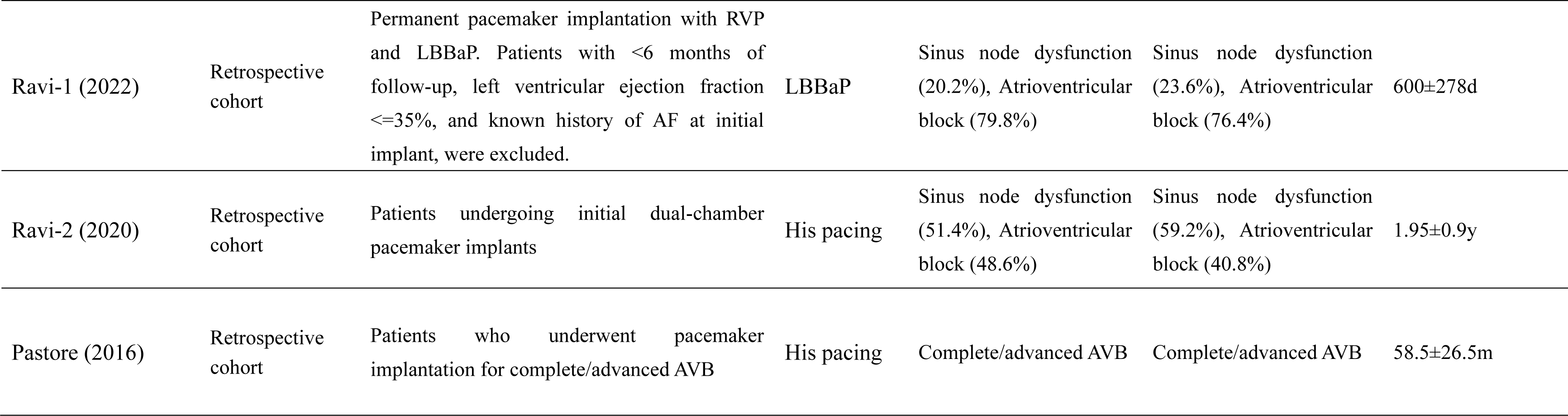
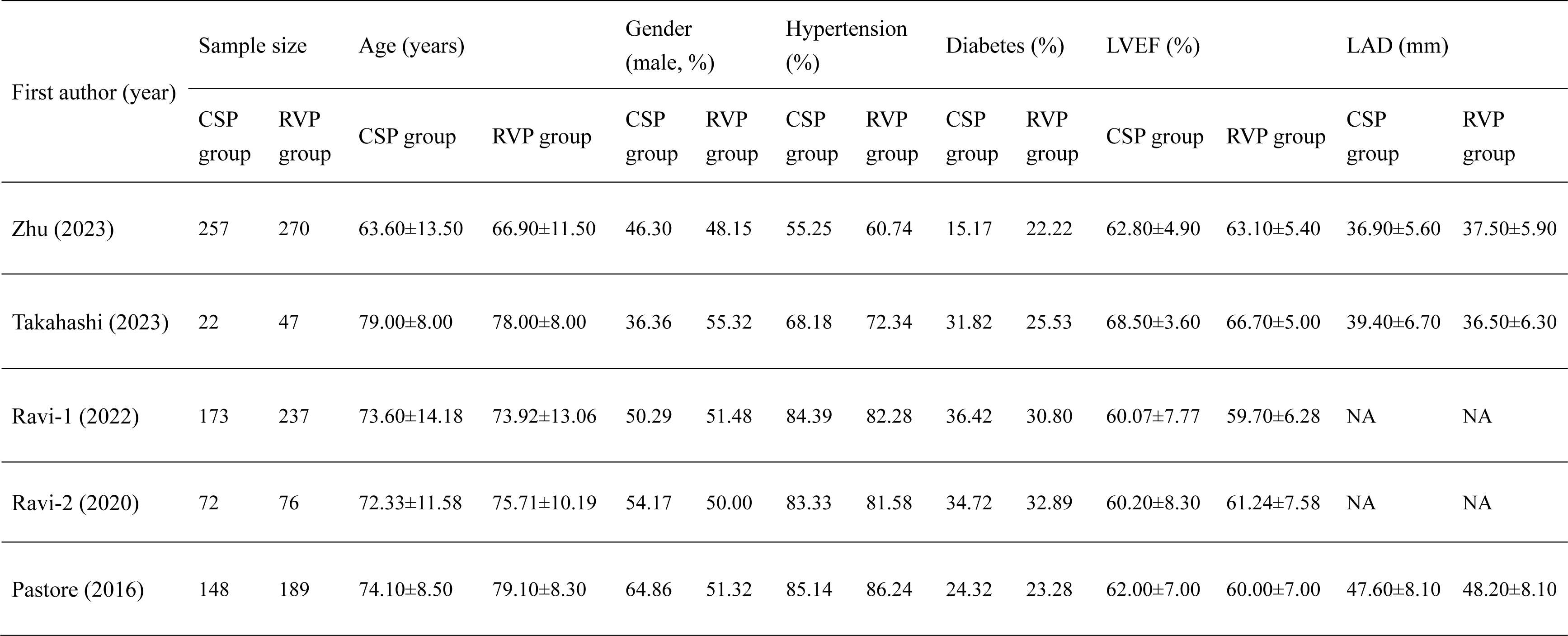
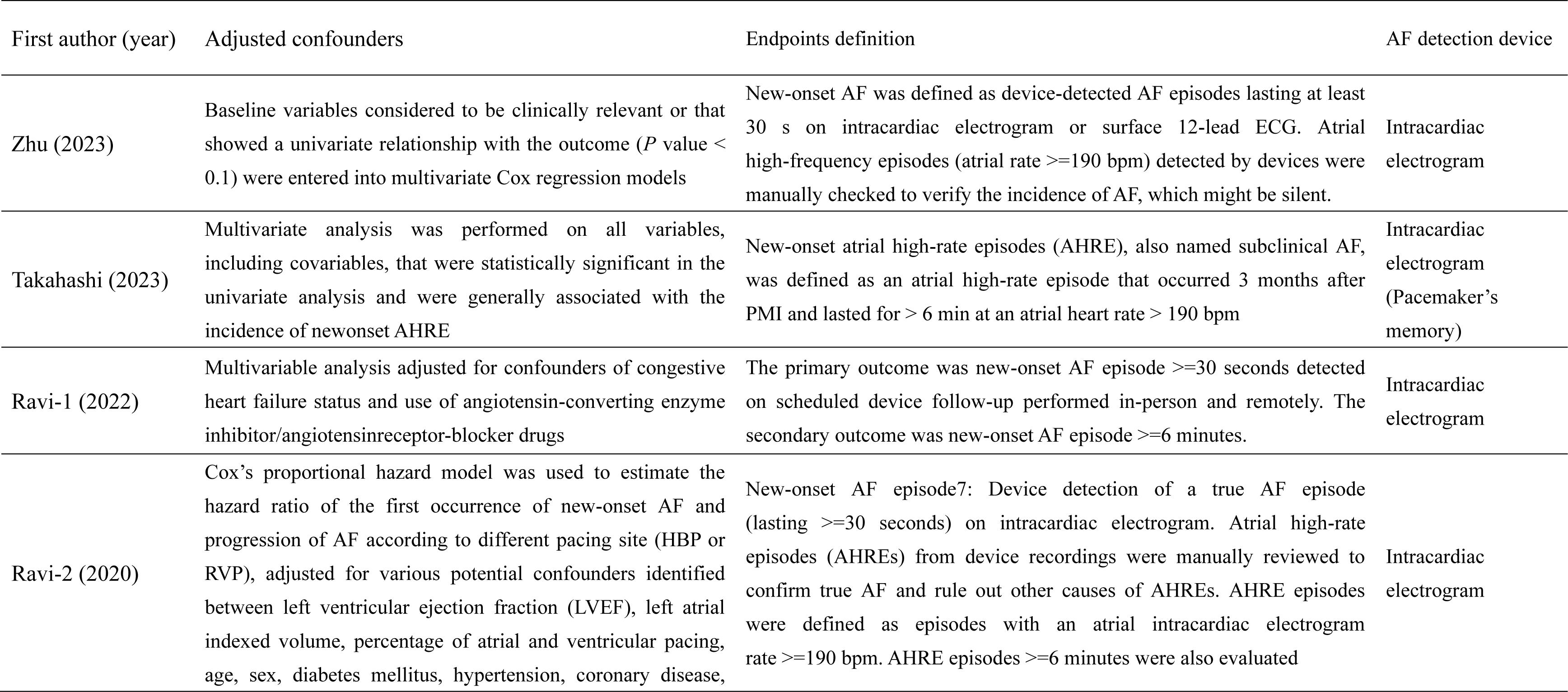
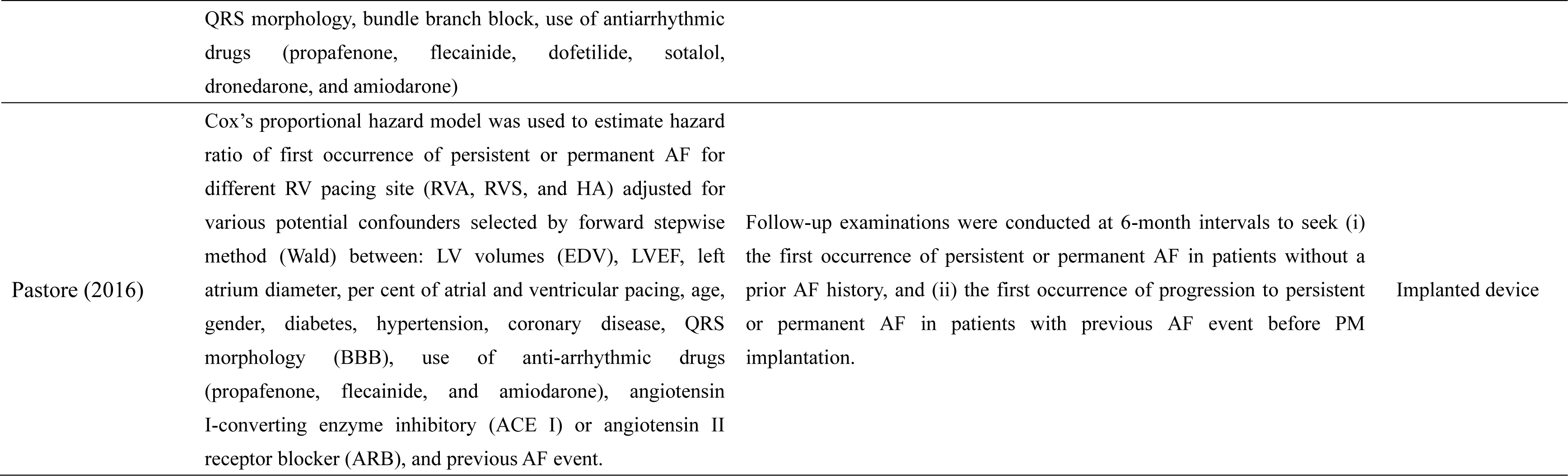
The baseline characteristics of eligible studies.

All five eligible studies[8–12] enrolled the patients requiring the pacemaker implantation (PMI), in which the indication of implant with sinus node dysfunction (SND) and atrioventricular block (AVB) was emphasized in three eligible studies[8, 10, 11], and that with only AVB was highlighted in two eligible studies[9, 12]. Two eligible studies[8, 10] reported LBBaP, and three[9, 11, 12] reported His pacing. New-onset AF was reported as clinical AF[8, 10–12] and subclinical AF (also named atrial high-rate episodes)[9]. Three studies reported the Vp with the cutoff value of 20% (including Vp ≥ 20%[8, 10, 11] and Vp < 20% subgroup[8, 10, 11]), and two studies reported with Vp with the cutoff value of 40% (including Vp ≥ 40%[8, 11] and Vp < 40% subgroup[8]). Only one study[12] compared the effects of three different pacing sites (such as His pacing, right ventricular apex, and right ventricular septal) on the new-onset AF, whereas the risk of new-onset AF between His pacing and right ventricular apecx pacing was available for further analysis in this study. The mean follow-up time from the eligible studies ranged from 11.1 months to 58.5 months[8–12]. All five observational studies showed a moderate-to-high quality (**Supplementary Table 1)**.

### 3.2 The pooled new-onset AF rates for CSP and RVP group

All eligible studies[8–12] reported the new-onset AF rate for CSP therapy and RVP therapy, respectively. The pooled rates of new-onset AF with random-effect model were 0.09 (95% CI, 0.05-0.14; *P* = 0.00; I^2^ = 68.54%; **Figure 2A**) and 0.26 (95% CI, 0.18-0.36; *P* = 0.00; I^2^ = 86.93%; **Figure 2B**), respectively, for CSP group and RVP group, with an interaction *P* value of 0.001. This result suggested that the rate of new-onset AF in CSP group was significantly lower compared with the RVP group.

**Figure 2.**
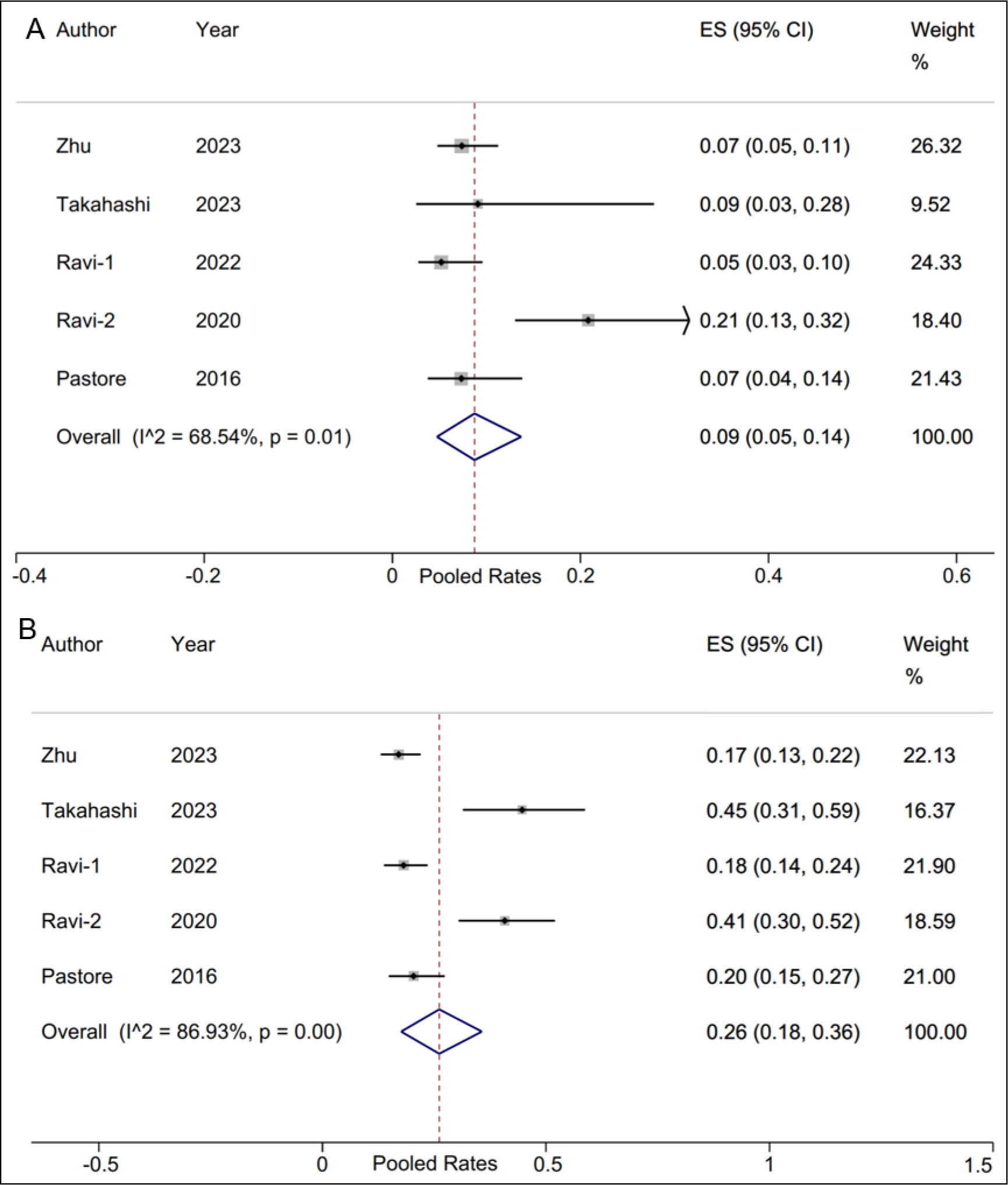
Forest plot of the new-onset AF rates for CSP and RVP group. A. The pooled rates of new-onset AF for CSP group; B. The pooled rates of new-onset AF for RVP group. CSP: conduction system pacing; RVP: right ventricular pacing; AF: atrial fibrillation.

Sensitivity analysis showed that no substantial change was identified in the overall combined proportion, which ranged from 0.07 (95% CI, 0.05-0.09) to 0.10 (95% CI, 0.06-0.19) for CSP group, and 0.23 (95% CI, 0.15-0.34) to 0.29 (95% CI, 0.18-0.45) for RVP group. This result revealed that no single study dominated the combined proportion and heterogeneity.

Subgroup analysis was performed and the results displayed in **Table 2** and **Table 3**. For the new-onset AF rate for CSP therapy, only one significant intervention-covariate interaction was identified in sample size subgroup, including > 100 (pooled rate 0.07; 95% CI, 0.05-0.09) and ≤ 100 (pooled rate 0.18; 95% CI, 0.10-0.26) with *P*=0.001 for interaction. Meanwhile, for the new-onset AF rate for RVP therapy, two significant intervention-covariate interactions were identified in sample size subgroup, including > 100 (pooled rate 0.18; 95% CI, 0.15-0.21) and ≤ 100 (pooled rate 0.42; 95% CI, 0.32-0.52) with *P* = 0.000 for interaction, as well as new-onset AF type subgroup, including clinical AF (pooled rate 0.23; 95% CI, 0.15-0.31) and subclinical AF (pooled rate 0.45; 95% CI, 0.31-0.59) with *P*=0.006 for interaction.

**Table 2.**
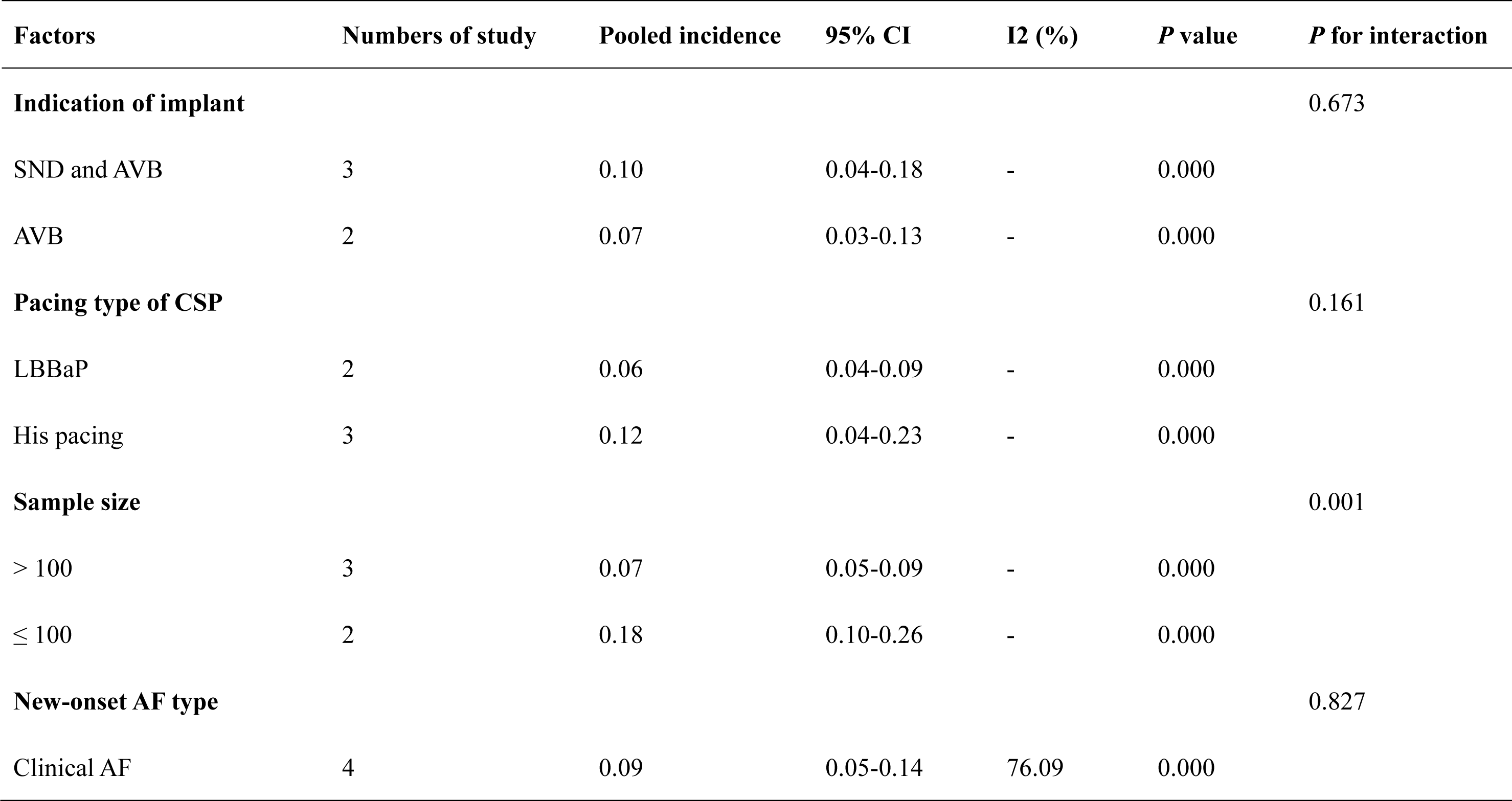
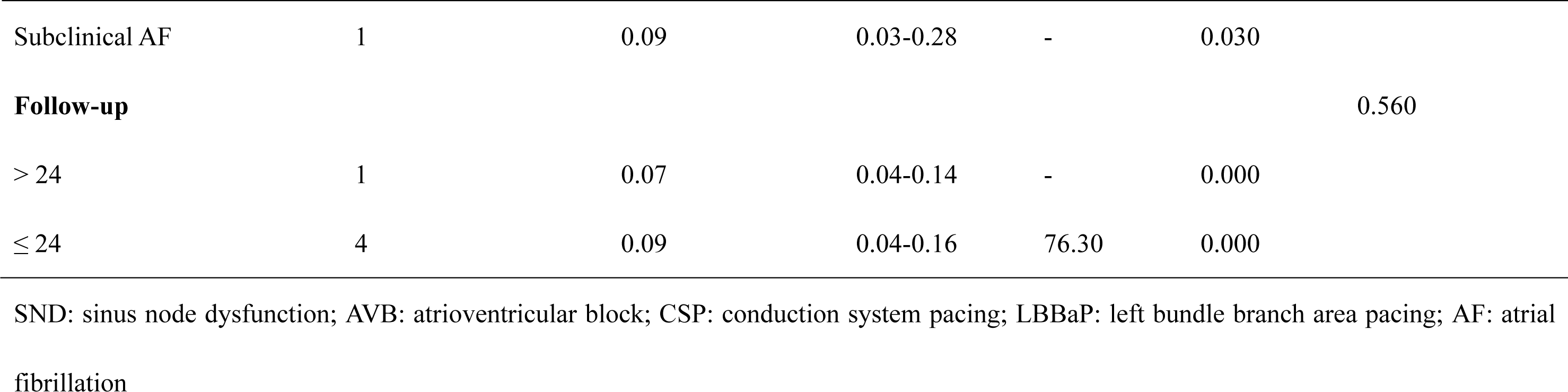
The subgroup analysis for the rate of new-onset AF for CSP group.

**Table 3.**
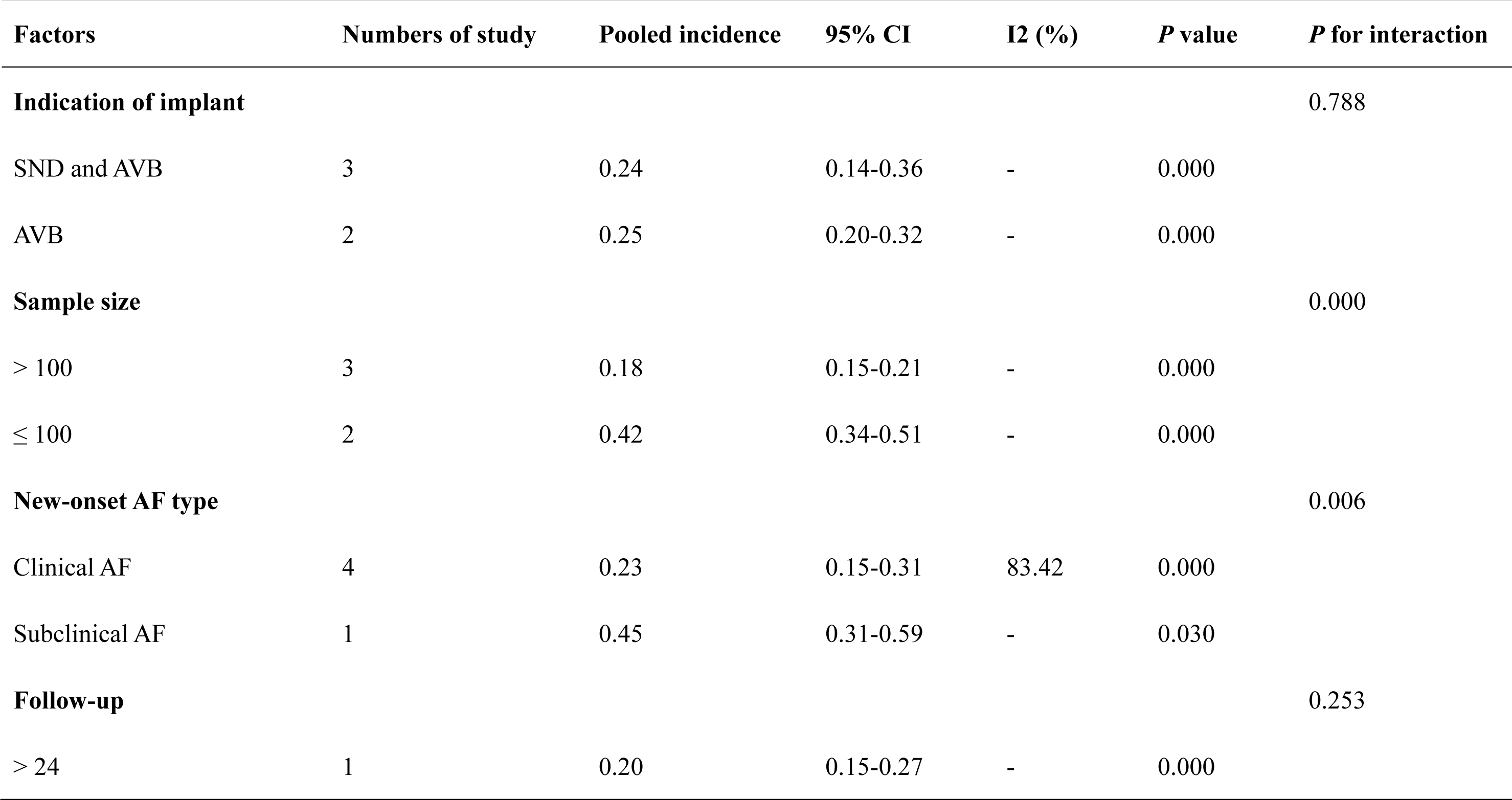
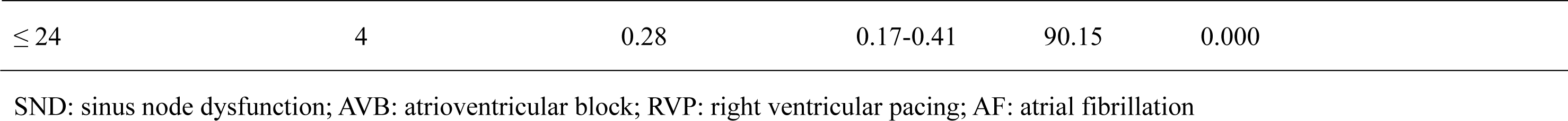
The subgroup analysis for the rate of new-onset AF for RVP group.

| Factors | Numbers of study | Pooled incidence | 95% CI | I<sup>2</sup> (%) | *P* value | *P* for interaction |
| --- | --- | --- | --- | --- | --- | --- |
| **Indication of implant** |  |  |  |  |  | 0.788 |
| SND and AVB | 3 | 0.24 | 0.14-0.36 | - | 0.000 |  |
| AVB | 2 | 0.25 | 0.20-0.32 | - | 0.000 |  |
| **Sample size** |  |  |  |  |  | 0.000 |
| > 100 | 3 | 0.18 | 0.15-0.21 | - | 0.000 |  |
| ≤ 100 | 2 | 0.42 | 0.34-0.51 | - | 0.000 |  |
| **New-onset AF type** |  |  |  |  |  | 0.006 |
| Clinical AF | 4 | 0.23 | 0.15-0.31 | 83.42 | 0.000 |  |
| Subclinical AF | 1 | 0.45 | 0.31-0.59 | - | 0.030 |  |
| **Follow-up** |  |  |  |  |  | 0.253 |
| > 24 | 1 | 0.20 | 0.15-0.27 | - | 0.000 |  |

|  |  |  |  |  |  |
| --- | --- | --- | --- | --- | --- |
| $\leq 24$ | 4 | 0.28 | 0.17-0.41 | 90.15 | 0.000 |

SND: sinus node dysfunction; AVB: atrioventricular block; RVP: right ventricular pacing; AF: atrial fibrillation

### 3.3 The risk of new-onset AF between CSP and RVP group

The risk of new-onset AF was assessed between CSP and RVP group from two aspects, including the pooled risk and adjusted risk. A total of five studies[8–12], including 633 patients with CSP therapy and 787 with RVP therapy in our study, reported the risk of new-onset AF between CSP group and RVP group. Compared with RVP group, CSP group was significantly associated with a lower new-onset AF risk (RR, 0.38; 95% CI, 0.28-0.51; *P* = 0.000; I^2^ = 0.00%; **Figure 3**) via a fixed-effect model.

**Figure 3.**
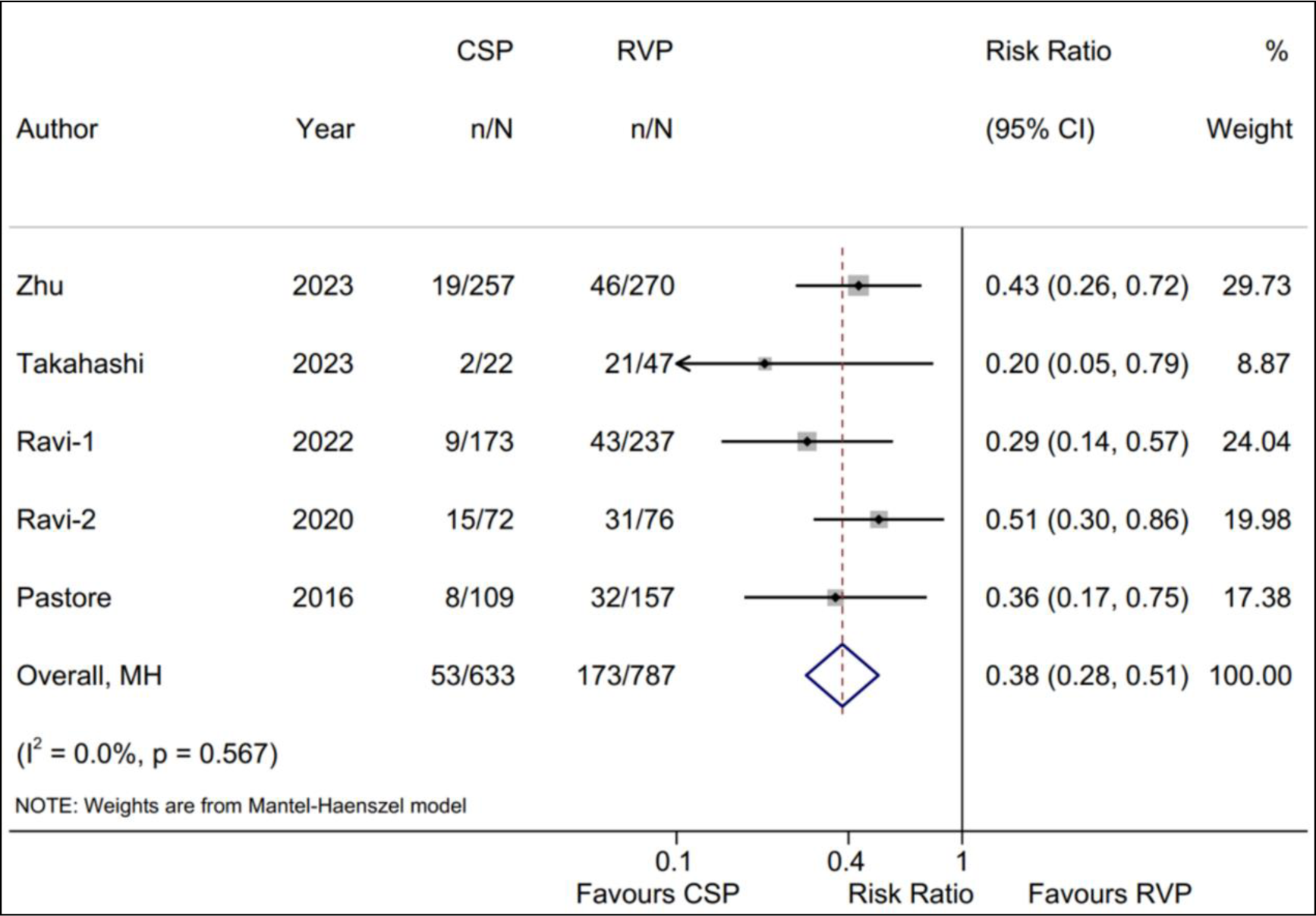
Forest plot of the risk of new-onset AF between CSP and RVP group. Comparison of the rate of new-onset AF between CSP and RVP group. CSP: conduction system pacing; RVP: right ventricular pacing; AF: atrial fibrillation.

We also performed sensitivity analysis for the pooled risk of new-onset AF and found that no significant change in the overall combined proportion, ranging from 0.35 (95% CI, 0.25-0.49) to 0.41 (95% CI, 0.30-0.57). This analysis revealed that the combined proportion and heterogeneity could not be dominated via a single study (**Supplementary Figure 1**). Meanwhile, publication bias was not displayed with Egger’s test (*P* = 0.079). These results suggested that our result was robust. The subgroup analysis for the pooled risk of new-onset AF was performed and the results were consistent with the pooled risk (**Table 4**).

**Table 4.** The subgroup analysis for the pooled risk of new-onset AF between CSP and RVP group.

| Factors | Numbers of study | Risk Ratio | 95% CI | I2 (%) | *P* value | *P* for interaction |
| --- | --- | --- | --- | --- | --- | --- |
| **Indication of implant** |  |  |  |  |  | 0.447 |
| SND and AVB | 3 | 0.41 | 0.29-0.56 | 0.00 | 0.000 |  |
| AVB | 2 | 0.31 | 0.16-0.59 | 0.00 | 0.000 |  |
| **Pacing type of CSP** |  |  |  |  |  | 0.811 |
| LBBaP | 2 | 0.37 | 0.24-0.55 | 0.00 | 0.000 |  |
| His pacing | 3 | 0.40 | 0.26-0.60 | 0.00 | 0.000 |  |
| **Sample size** |  |  |  |  |  | 0.682 |
| > 100 | 3 | 0.37 | 0.26-0.52 | 0.00 | 0.000 |  |
| ≤ 100 | 2 | 0.42 | 0.25-0.68 | 39.30 | 0.001 |  |
| **New-onset AF type** |  |  |  |  |  | 0.345 |
| Clinical AF | 4 | 0.40 | 0.30-0.54 | 0.00 | 0.000 |  |
| Subclinical AF | 1 | 0.20 | 0.05-0.79 | 0.00 | 0.022 |  |
| **Follow-up** |  |  |  |  |  | 0.870 |
| > 24 | 1 | 0.36 | 0.17-0.75 | 0.00 | 0.006 |  |
| ≤ 24 | 4 | 0.38 | 0.28-0.53 | 0.00 | 0.000 |  |

SND: sinus node dysfunction; AVB: atrioventricular block; CSP: conduction system pacing; RVP: right ventricular pacing; LBBaP: left bundle branch area pacing; AF: atrial fibrillation

In addition, the adjusted risk of new-onset AF was reported by five eligible studies[8–12]. Similarly, we found that compared with RVP group, CSP group was significantly associated with a lower adjusted risk of new-onset AF (HR, 0.33; 95% CI, 0.24-0.44; *P* = 0.000; I^2^ = 0.00%; **Figure 4**) via a fixed-effect model. Sensitivity analysis also indicated that that no significant change, ranging from 0.28 (95% CI, 0.20-0.40) to 0.35 (95% CI, 0.24-0.50), in the overall combined proportion. Publication bias using Egger’s test (*P* = 0.768) was not displayed. The subgroup analysis for the adjusted risk of new-onset AF showed the consistent results with the pooled adjusted risk (**Figure 5**).

**Figure 4.**
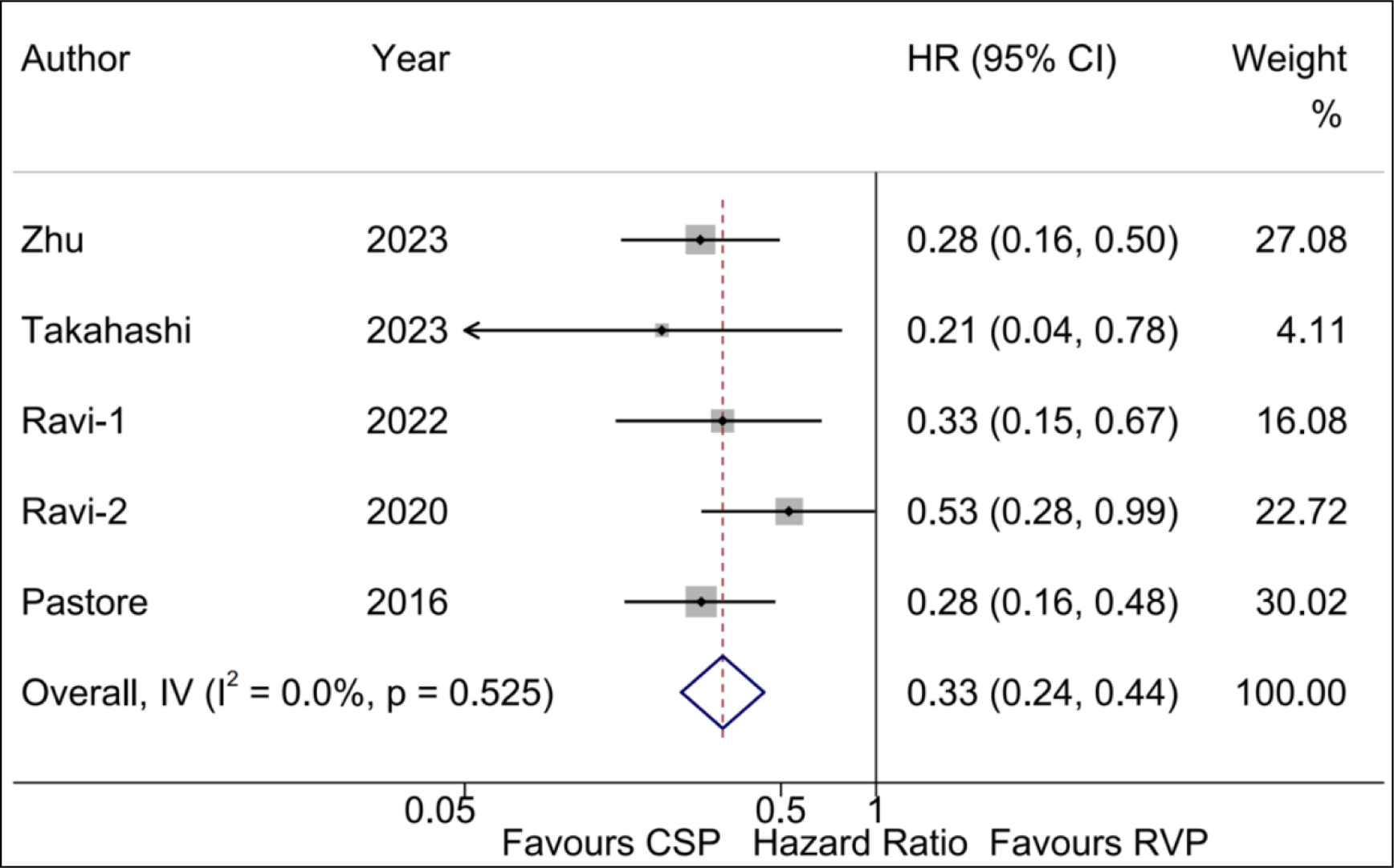
Forest plot of the adjusted risk of new-onset AF between CSP and RVP group. Comparison of the adjusted risk of new-onset AF between CSP and RVP group. CSP: conduction system pacing; RVP: right ventricular pacing; AF: atrial fibrillation.

**Figure 5.**
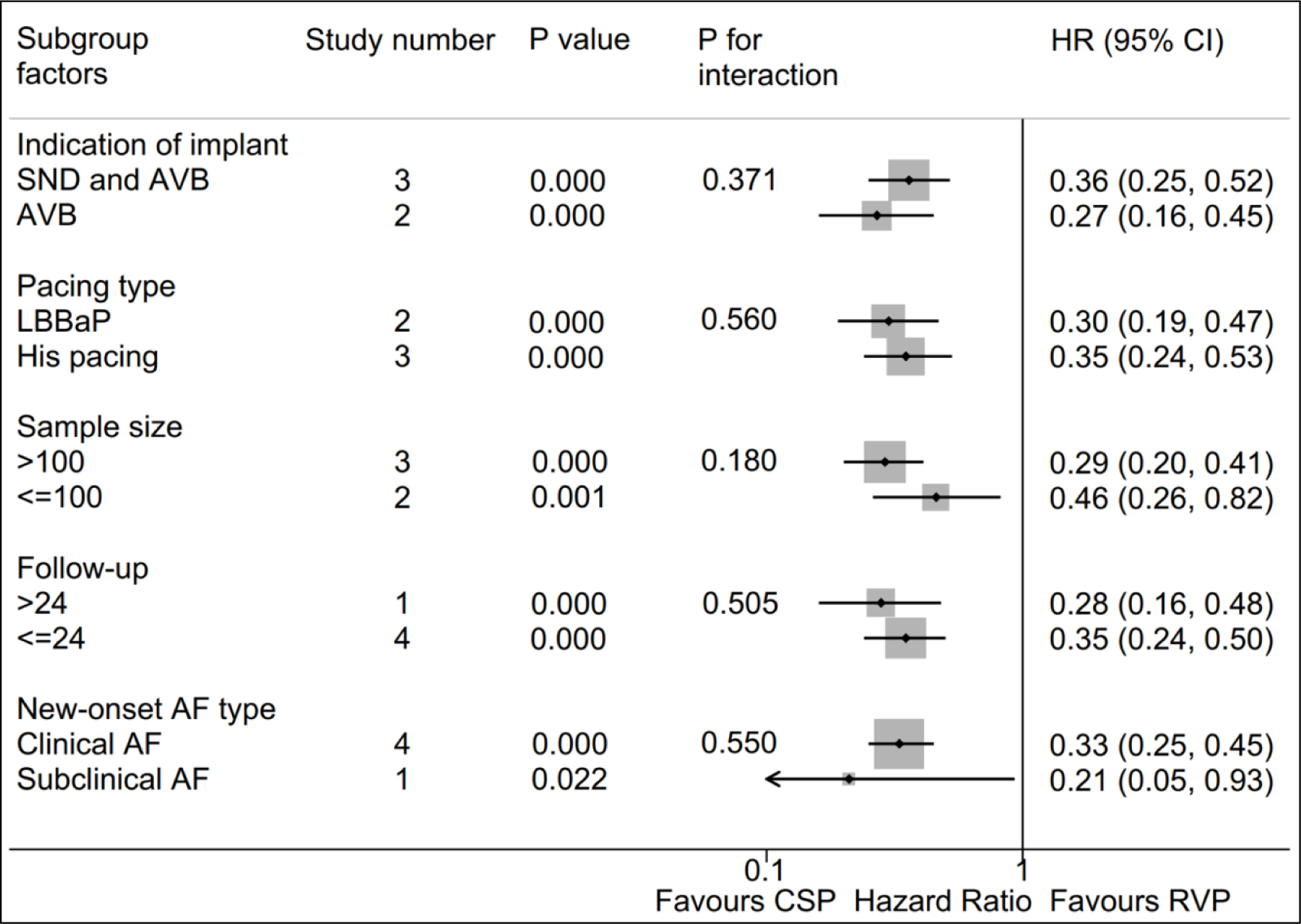
Forest plot of the adjusted risk of new-onset AF between CSP and RVP group. Subgroup analysis of the adjusted risk of new-onset AF between CSP and RVP group. CSP: conduction system pacing; RVP: right ventricular pacing; AF: atrial fibrillation.

### 3.4 The adjusted new-onset AF risk between CSP and RVP group based on Vp

Three eligible studies[8, 10, 11] reported the adjusted risk of new-onset AF between CSP and RVP group based on Vp ≥ 20% and Vp < 20%. The pooled HRs for the adjusted new-onset AF risk between CSP and RVP group with Vp ≥ 20% and Vp < 20% were 0.24 (95% CI, 0.16–0.37; *P* = 0.000) and 0.65 (95% CI, 0.35–1.21; *P* = 0.000), respectively. Importantly, a statistically significant intervention-covariate interaction was identified in Vp subgroup, including Vp ≥ 20% and Vp < 20% with *P* = 0.010 for interaction (**Figure 6**).

**Figure 6.**
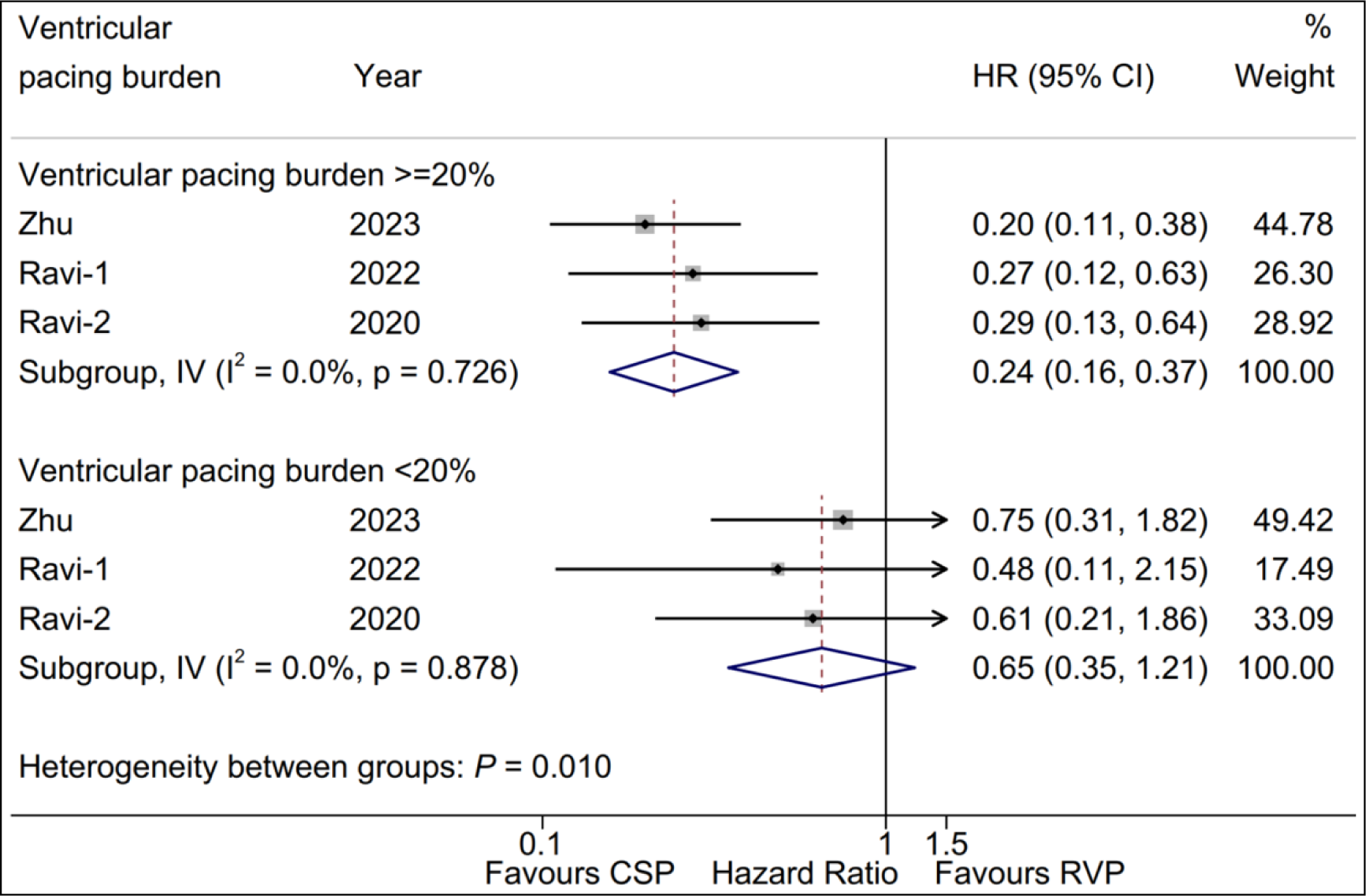
Forest plot of the adjusted risk of new-onset AF between CSP and RVP group based on Vp ≥ 20% subgroup and Vp < 20% subgroup. CSP: conduction system pacing; RVP: right ventricular pacing; AF: atrial fibrillation; Vp: ventricular pacing burden.

Also, we evaluated the adjusted risk of new-onset AF between CSP and RVP group based on Vp ≥ 40% and Vp < 40% with two eligible studies[8, 11]. We found that compared with RVP group, the adjusted new-onset AF risk of CSP group was significantly decreased in Vp ≥ 40% subgroup (HR = 0.25; 95% CI, 0.14–0.42; *P* = 0.000) and it was not in Vp < 40% subgroup (HR = 0.57; 95% CI, 0.26–1.27; *P* = 0.168), respectively. Additonally, no statistically significant intervention-covariate interaction was displayed between Vp subgroups with *P* = 0.088 for interaction (**Figure 7**).

**Figure 7.**
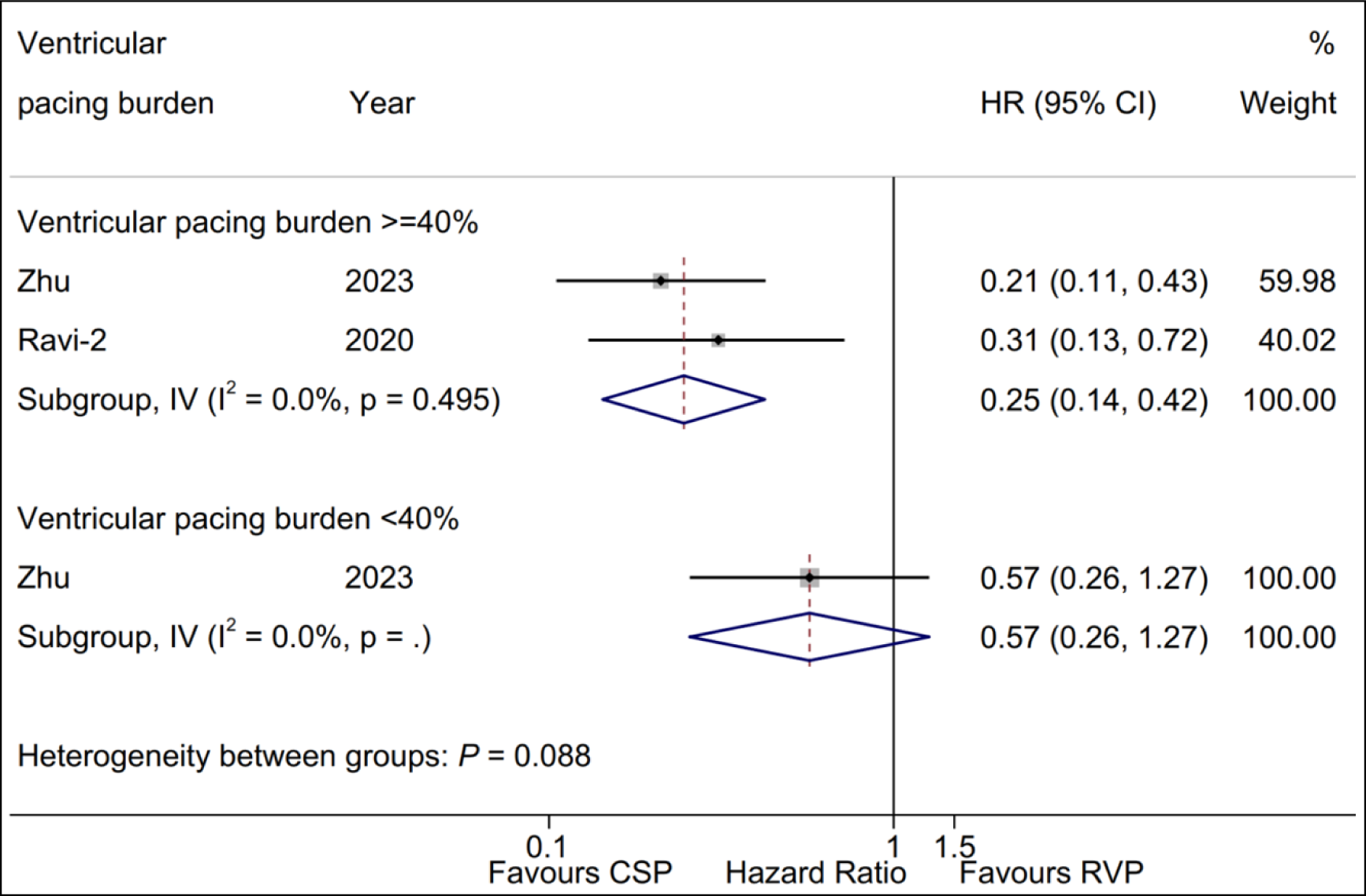
Forest plot of the adjusted risk of new-onset AF between CSP and RVP group based on Vp ≥ 40% subgroup and Vp < 40% subgroup. CSP: conduction system pacing; RVP: right ventricular pacing; AF: atrial fibrillation; Vp: ventricular pacing burden.

## 4. Discussion

We comprehensively enrolled five observational studies with 1,491 patients requiring pacing therapy, including 672 patients in CSP group and 819 patients in RVP group. To our knowledge, our study may be the first registered meta-analysis to compare the risk of new-onset AF between CSP and RVP group. The main findings included: 1) The rate of new-onset AF in CSP group was significantly lower in comparison with the RVP group; 2) Compared with RVP group, CSP group showed a lower pooled risk, as well as the adjusted risk of new-onset AF; 3) A statistically significant intervention-covariate interaction with *P* = 0.010 was identified in Vp subgroup, including Vp ≥ 20% and Vp < 20%. The schematic representation for this study is displayed in **Figure 8**.

**Figure 8.**
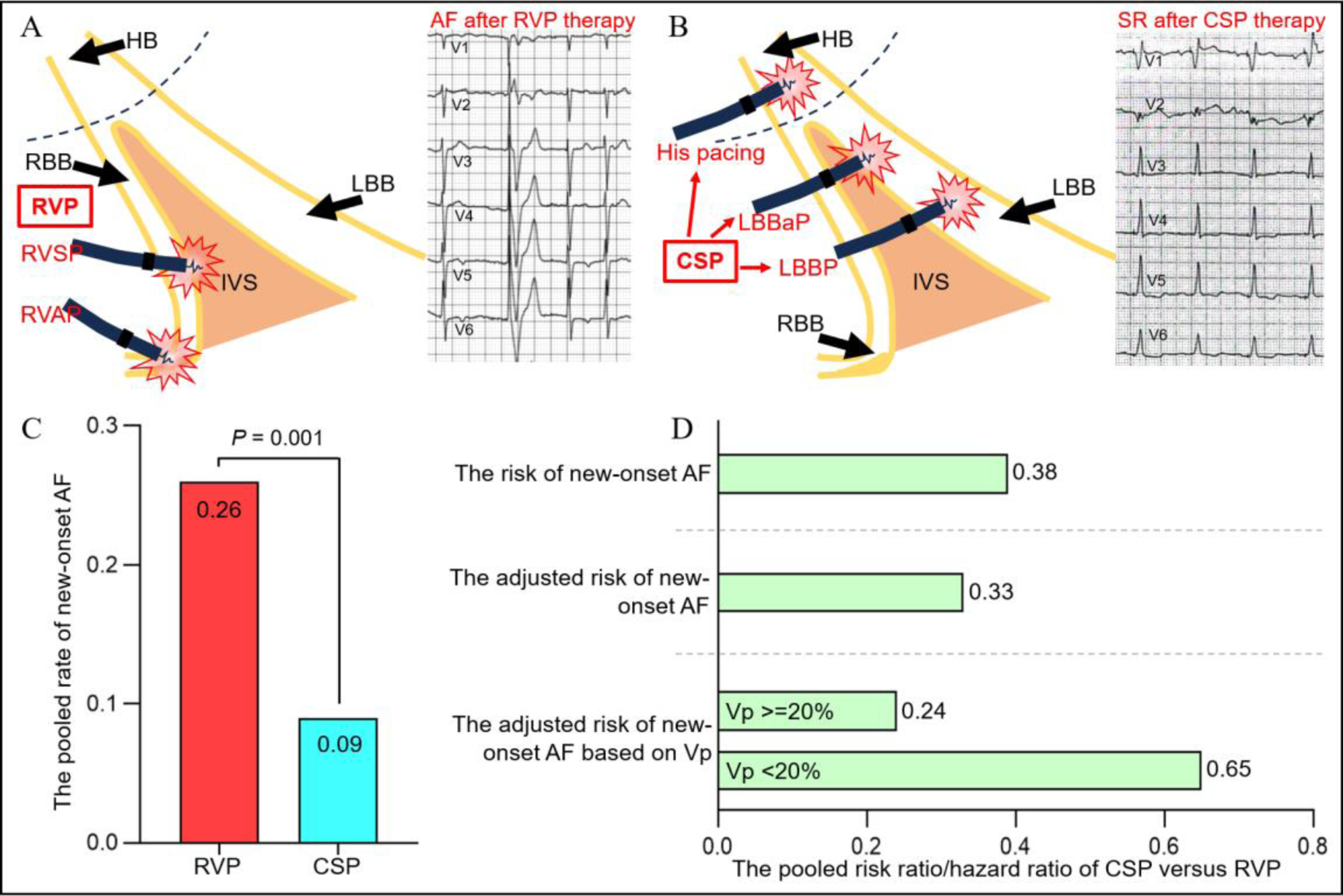
Schematic Representation. A. Pacing pattern diagram and related electrocardiogram for RVP; B. Pacing pattern diagram and related electrocardiogram for CSP; C: The histogram for the new-onset AF rates for CSP and RVP group; D. The new-onset AF risk ratio or hazard ratio between CSP and RVP. CSP: conduction system pacing; RVP: right ventricular pacing; HB: His bundle; RBB: right bundle branch; LBB: left bundle branch; AF: atrial fibrillation; SR: sinus rhythm; IVS: interventricular septum; RVAP: right ventricular apex pacing; RVSP: right ventricular septal pacing; LBBP: left bundle branch pacing; LBBaP: left bundle branch area pacing; Vp: ventricular pacing burden.

RVP, a well-established pacing strategy, has been widely utilized for patients requiring pacing therapy due to relatively easy implantation, satisfying efficacy, and peocedural safety. Whereas, available evidence suggests that RVP is significantly related to the ventricular electricity and contraction dyssynchrony, promoting to the occurrence of pacing induced cardiomyopathy, ultimately leading to increased risk for atrial arrhythmias, heart failure, and mortality[13]. New-onset AF, including clinical and subclinical AF, is of concern during pacing therapy, considering the risks of progressing into persistent atrial fibrillation, increasing rate of stroke/systolic embolism, left ventricular dysfunction, and impaired quality of life[14]. Numerous studies had demonstrated that RVP was significantly associated with an increasing risk of new-onset AF, especially with the patients requring the high proportion of ventricular pacing[15–18]. Therefore, an alternative pacing strategy is needed to decrease the risk of new-onset AF after pacing therapy.

CSP is an emerging and intriguing physiological pacing modality, facilitating decrease, or even reverse the adverse clinical outcomes associated with RVP. Our previuous study also revealed that CSP was more effective performance than conventional pacing therapy in heart failure patients, including shortened QRS duration, improved ventricular ejection fraction, improved NYHA class, higher response rate, as well as lower heart failure rehospitalization[7]. Meanwhile, an increasing evidence indicated that CSP was statistically associated with a reduced risk of new-onset AF in comparison of RVP[8, 10, 11]. In this study, our results also suggested that compared with RVP therapy, CSP therapy could significantly decrease the new-onset AF rate for patients requiring the pacing therapy, which might further increase evidence to highlight the superior perfomance on CSP.

Several risk factors have been reported to be involved in pacing induced pacing induced cardiomyopathy. Lee *et al*.[19] performed a retrospective study of 234 patients underwent a permanent pacemaker implantation with RVAP from 1982 to 2004, and found that older age at implantation was an idependent predictor for pacing induced heart failure (HR = 1.62; 95% CI, 1.22–2.16; *P* = 0.001). The latest meta-analysis performed by Somma *et al*.[20] with twenty-six studies including a total of 57,993 patients suggested that male gender, chronice kidney disease, previous myocardial infarction, native QRS duration, and paced QRS duration were identified to be the most important risk factors for pacing induced cardiomyopathy. Moreover, our previous studies also revealed that high-volume center (also meant a high sample size for a key technique) representing a relatively advanced operational team, was significantly associated with a better effacacy and comparable safety[7, 21]. Whereas, very few studies have reported the potential risk factors for the potential pacing induced new-onset AF. In this report, consistent with the previous studies[7, 21], we found that the sample size > 100 was considered as a key factor for reduction of the new-onset AF rate for CSP and RVP, respectively, in comparison of that less than 100. This results also provided an inspiration to our future research that the sample size with cutoff 100 might be appropriate and necessary to acquire the relatively stable and real-world rate, instead of the pseudo-high rate, of new-onset AF post pacing. Subclinical AF has been demonstrated to be associated with the risk of clinical AF[22]. A recent meta-analysis with eleven studies conducted by Mahajan R *et al*.[23] showed that the risk of clinical AF was as high as 5.7 (95% CI, 4.00–8.00; *P* < 0.001) in patients with subclinical AF. Our subgroup analysis for the subclinical AF rate with RVP therapy was significantly higher than clinical AF rate, whereas both rates were similar with CSP therapy. This result might partly explain the high new-onset AF rate in RVP therapy owing to the high incidence of subclinical AF.

Our study also indicated that CSP group could significantly reduce the pooled risk, as well as adjusted risk of new-onset AF risk compared with RVP group. Additionally, subgroup analysis showed similar results. While His pacing has been demonstrated to be the feasibility and safety of intrinsic conduction system pacing, whereas it failed to widespread performed due to multiple concerns, such as unstable and elevated thresholds and relatively with follow-up. Excitingly, LBBaP or LBBP has been more widely adopted relatively low and stable pacing thresholds[24, 25]. Interestingly, our study showed that the rate and risk of new-onset AF between LBBaP subgroup and His pacing subgroup were similar, which indicated a consistent effect between both types of CSP. However, concerns must be paid for the higher pacing threshold with His pacing during long-term follow-up, potentially leading to excessive battery consumption and premature pacing lead revision. In addition, our study also suggested that follow-up did not affect the role of CSP in reducing the new-onset AF.

Previous studies had demonstrated that individual with SND showed a high risk of new-onset AF on account of abnormal sinoatrial node function and increased automaticity of atrium[26]. A two-community based cohort study including a total of 19,893 individuals with a follow-up of seventeen years revealed that SND could significantly increase the risk of AF as high as 5.75 after adjustment of multiple cardiovascular diseases[27]. A latest meta-analysis with thirty-five studies performed by Liu *et al*.[28] demonstrated that patients with SND showed a higher new-onset AF risk in comparation of those with AVB (RR, 3.45; 95% CI, 1.54-7.69; *P* = 0.003). Therefore, we made a reasonable speculation that the new-onset AF rate, theoretically, should be higher in eligible studies with the indication of SND and AVB, while that be lower in these with indication of AVB. Whereas, in our study the similar results between SND and AVB subgroup and AVB subgroup were shown in the analysis of new-onset AF rates of CSP therapy and RVP therapy, as well as the risk of new-onset AF between CSP and RVP group. This result seemed contradictory to previous studies and our speculations. The potential explanations for our result are as follow: The pacing therapy with timely restoration of heart rates with normal atrioventricular conduction might significantly interrupt the natural progression of the SND and AVB, ultimately leading to the occurrence of new-onset AF that mainly depended on pacing percentage or pacing modality.

Vp was a key determinant in the new-onset AF risk between CSP and RVP group, with a positive linear relationship between the risk of AF and Vp[15]. Importantly, screening an optimal Vp cutoff value for recommendation of CSP therapy to prevent new-onset AF or even pacing induced cardiomyopathy is of great interes to the pacing community. The 2018 ACC/AHA/HRS guideline on the evaluation and management of patients with bradycardia and cardiac conduction delay suggested that more physiologic ventricular activation (such as His pacing) should be preferentially provided with the patients requiring Vp > 40%[29]. The lastest HRS/APHRS/LAHRS guideline defined the substantial Vp of ≥ 20%-40% and highlighted that patients who underwent pacemaker implantation and were expected to require substantial Vp may be considered for CSP to reduce pacing-induced cardiomyopathy risk[30]. Our study showed that CSP could significantly reduce the new-onset AF risk in Vp ≥ 20% subgroup, as well as Vp ≥ 40% subgroup, compared with RVP. Whereas, only one significant intervention-covariate interaction was identified between Vp ≥ 20% and Vp < 20% subgroup, rather than Vp ≥ 40% and Vp < 40% subgroup. The potential explanation might be that some patients (e.g., Vp 20%-40%) could benefit from CSP compared with RVP in Vp < 40% subgroup, which leading to a more overlap in 95%CI and no significant interaction between Vp ≥ 40% subgroup and Vp < 40% subgroup. More studies should be designed and performed to identify the minimal Vp cutoff value during 20%-40%, with that a significant interaction could be achieved, thus providing strong evidence for the guideline with optimal Vp cutoff value.

## 5. Limitations

There are several limitations in our study. First, all eligible studies in this analysis belonged to retrospective or observational studies, which has inherent limitations (such as inevitable confounding factors), thus leading to possiblely unreliable results[31]. Whereas, the adjusted risk of new-onset AF was analyzed to minimize the effect of confounders as much as possible. Notably, the pooled risk and the adjusted risk of new-onset AF from our eligible studies were similar, suggesting that our results were reliable. Our findings should be further assessed in further multicenter randomized trials with longer follow-up and larger sample size. Second, accumulated studies suggested that LBBP achieved left ventricular synchrony but induced right ventricular conduction delay[32]. Bipolar LBBP was expected to improve the bi-ventricular electrical synchrony in comparison of unipolar LBBP, futher achieving more physiological conduction system activation[33]. However, no data to date was reported on the risk of new-onset AF between bipolar and unipolar LBBP. Therefore, this might be a promising and novel perspective to provide related evidence for the optimal pacing therapy. Third, some potential biases derived from meta-analysis, might power our results. Considering that, sensitivity analysis and publication bias test both suggested our results were relatively robust.

## 6. Conclusions

Our study suggests that CSP is superior to reduce the new-onset atrial fibrillation risk compared with RVP. The Vp ≥ 20% may be the key determinant of the lower risk of new-onset AF with CSP therapy. More multicenter randomized trials with longer follow-up and larger sample size should be conducted to further confirm our findings.

## Data Availability

The data that support the findings of this study are available from the corresponding author upon reasonable request.

## Acknowledgments

We sincerely appreciated the supporting by the Suzhou Science and Technology Plan Project (SKY2022151). We would also show sincere appreciation to the reviewers for critical comments on this article.

---

**Supplementary Figure 1.**
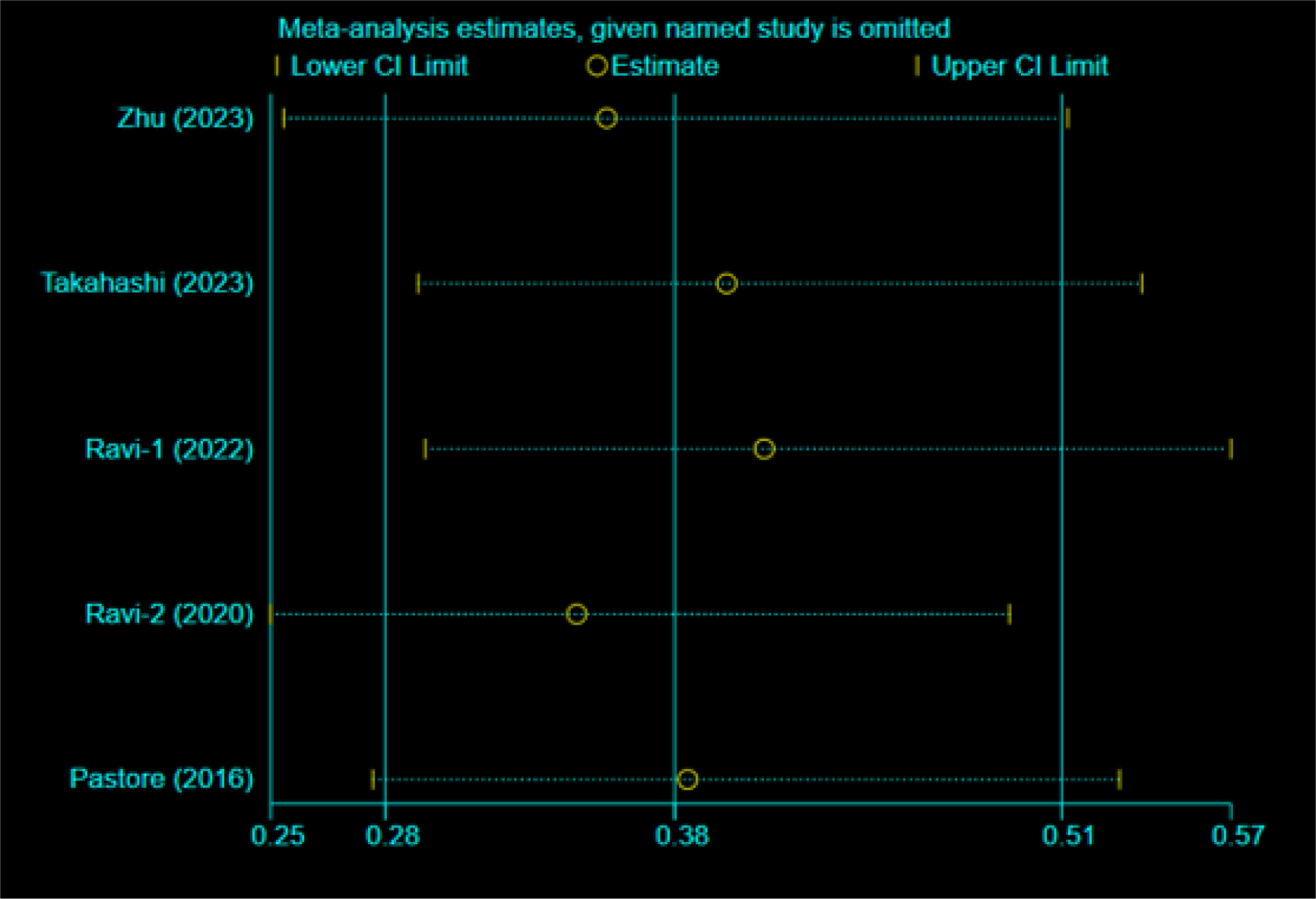
The sensitivity analysis for the pooled risk of new-onset AF.

